# Depression polygenic scores are associated with major depressive disorder diagnosis and depressive episode in Mexican adolescents

**DOI:** 10.1101/2020.05.31.20118711

**Authors:** Jill A. Rabinowitz, Adrian I. Campos, Corina Benjet, Jinni Su, Luis Macias-Kauffer, Enrique Méndez, Gabriela A. Martinez-Levy, Carlos S. Cruz-Fuentes, Miguel E. Rentería

## Abstract

**Objective:** Large-scale genome-wide association studies have uncovered genetic variants associated with depression; however, most of this work has been limited to adults of European ancestry. We investigate the ability of depression polygenic risk scores (PRS) to predict both lifetime and past year major depressive disorder (MDD) diagnosis and the experience of a major depressive episode (MDE) in a sample of adolescents with admixed ancestry from Mexico City, and explore whether adverse life events moderate these relations.

**Methods:** The study sample consisted of adolescents (N = 1,152) aged 12–17 from Mexico City who were interviewed and genotyped as part of a general population survey on adolescent mental health. PRS for depression were derived using summary statistics from a large-scale discovery genome-wide association study (GWAS) conducted on depressive symptoms that included over 800,000 individuals of European ancestry (Howard et al., 2019).

**Results:** Higher depression PRS were associated with a higher likelihood of both past year MDD and MDE and lifetime MDE, accounting for 1.5%-2.5% of the variance in these outcomes. Adversity did not moderate the relationship between depression PRS and lifetime or past year MDD or MDE.

**Limitations:** This study is cross-sectional. As such, a few participants might have experience MDD/MDE after the interview. In addition, our sample comprised only Mexican youth and thus it may not generalize to other populations.

**Conclusion:** Our results indicate that depression PRS derived from a European ancestry GWAS are associated with MDD and MDE risk among Mexican adolescents and have the potential to aid in the identification of youth who may be genetically prone to developing depressive symptoms.

## Introduction

Depression is a highly heterogeneous and complex disorder. Around the world, it affects over 350 million people and is one of the leading causes of disability. It is often comorbid with other psychiatric conditions (e.g., anxiety) (Costello et al., 2003), chronic physical diseases (e.g., diabetes) (Jokela et al., 2019), lower educational attainment (Fletcher, 2010), and increased mortality (Chiu et al., 2018). Although depression can occur at any time during the developmental course, depression onset during adolescence is quite common, with close to 10% of adolescents reporting a depressive episode before the age of 21 (Mojtabai et al., 2016). Relative to adult-onset depression, depression with onset in adolescence often persists throughout the life course and is associated with higher rates of morbidity and mortality (Thompson, 2008; Wilson et al., 2015).

A substantial body of work indicates that depression is genetically influenced, with twin studies indicating a moderate heritability (46% for adults aged 50–92; 40% for younger adults aged 23–28) (Corfield et al., 2017). Although several candidate genes have been linked to major depression (e.g., serotonin transporter polymorphism) (Chipman et al., 2007), these studies have often failed to replicate, dampening the enthusiasm among researchers and practitioners alike in incorporating these candidate genes in precision medicine initiatives and clinical care (Border et al., 2019). More recently, a number of large-scale genome-wide association studies (GWAS) have been conducted to identify the genetic architecture of depression (Howard et al., 2019; Okbay et al., 2016). The increase in GWAS is based on the premise that psychiatric disorders are not a byproduct of single genes, but rather the result of complex interactions between hundreds of thousands of genetic variants (Maher, 2015). The most recent GWAS on depression, which included a total sample of over 800 thousand participants, yielded 102 independent genetic variants associated with depression. Eighty-seven of those variants were replicated in an independent sample of more than 1.3 million people (Howard et al., 2019). In the same study, a depression polygenic risk score (PRS) explained between 1.5%-3.2% of the variance in phenotypic depression risk in clinically ascertained samples (Howard et al., 2019).

To date, most GWAS analyses have been almost entirely limited to individuals of European descent. The extent to which findings from these studies apply to different ancestry groups needs to be examined given population-specific genetic variation (Martin et al., 2018). Extant work has indicated that depression PRS were associated with childhood-persistent depression and early-adult onset depression in a predominantly European-ancestry sample (Kwong et al., 2019). Other work has shown that a depression PRS was associated with increasing and high depressive symptoms across childhood and adolescence (Lussier et al., 2020) and a steep increase in depressive symptoms in adolescence and young adulthood (Kwong et al., 2020) in European-ancestry samples. Additional research has indicated that in a European-ancestry sample, depression PRS were associated with late adolescent depression, but not low or early-onset adolescent depression (Rice et al., 2019). In sum, research investigating the relationship between depression genetic liability and phenotypic depression has been conducted disproportionately among adults of European ancestry; thus, work examining these relationships in other populations at different developmental periods is wanting.

A particular population that may benefit from work investigating the relationship between depression genetic liability and phenotypic depression is Mexican youth. In a previous study, close to 13% of adolescents in Mexico City’s metropolitan area met criteria for a lifetime diagnosis of MDD in adolescence with a reoccurrence rate of close to 50% in young adulthood (Benjet et al., 2019). Given that Mexican youth may be at increased risk for depression and the adverse sequelae associated with this condition, an examination of individual-specific (i.e., genetic) and contextual contributors to this disorder is warranted to inform interventions aimed at reducing depression onset, improving course and treatment responsiveness.

In addition to genetic differences that might confer risk for depression, the environmental context can significantly affect the extent to which this occurs. Consistent with ecological systems and developmental psychopathology theories, many interacting forces between biological, environmental, and developmental systems influence depression vulnerability (Starr et al., 2014). One well-documented environmental contributor to depression is adverse life events (Llabre et al., 2017). For example, experiencing adversity (i.e., sexual abuse, having a parent with depression, family disruption) during childhood or adolescence has been associated with a depression diagnosis among Mexican (Benjet et al., 2019) and Latino adolescents (Jaschek et al., 2016). Moreover, exposure to adversity may profoundly shape the developing brain and neurobiological systems that make some individuals more sensitive to future stressors (McIlwricka et al., 2016; Starr et al., 2014). Moreover, exposure to higher levels of adversity may also interfere with youth meeting developmentally salient demands across development(i.e., academic success, employment), exacerbating risk for the development of depressive symptoms in adulthood (Starr et al., 2014).

The examination of putative early adversity x candidate gene interactions in predicting depression are plenty, with several interactions between early adversity and candidate genes among European-ancestry young adults having been documented (Starr et al., 2014; Taylor et al., 2006). However, a recent meta-analysis was unable to replicate previously identified candidate gene x environment moderation effects related to depression (Border et al., 2019). Although the consideration of polygenic influences is considered the state of the science and adversity has been strongly linked to depression, there is a scarcity of work that has examined the joint contributions of these two factors. Available work revealed that a composite index of childhood trauma was associated with heightened risk for depression among individuals with *lower* polygenic depression risk, although these findings were not replicated in an independent sample (Mullins et al., 2016). Other work has found that adversity interacted with genome-wide single nucleotide polymorphisms to predict depression among women of Chinese ancestry (Peterson et al., 2018).

To our knowledge, no work has examined whether depression could be predicted from the interplay between adversity and depression polygenic risk among Mexican youth; such information could elucidate what contributes to the etiology, course, and persistence of depressive symptoms in this population. The current study leverages the results of the most extensive genetic discovery efforts to date on depression, to investigate whether (a) depression PRS are associated with lifetime and past year MDD diagnosis or MDE, and (b) adversity moderates the relationship between depression genetic liability and MDD or MDE in a sample of Mexican adolescents. This work is consistent with a recent call for work to consider the joint contributions of context and biology among ethnic minority populations (Causadias and Cicchetti, 2018).

## Methods

### Participants

Individuals in the present study were participants in an adolescent mental health population survey conducted by Mexico’s National Institute of Psychiatry “Ramón de la Fuente Muñíz” in 2005 (Benjet et al., 2019). Three thousand and five individuals between the ages of 12 and 17 living in Mexico City were interviewed face-to-face in their homes by trained personnel using the computer-assisted World Mental Health Composite International Diagnostic Interview for Adolescents (WMH-CIDI-A) (Merikangas et al., 2010). This fully structured interview generates psychiatric diagnoses based on DSM-IV criteria. The sampling was designed by expert epidemiologists to be representative of the population of Mexico City and surrounding municipalities. The final sample size with both genetic and phenotypic data after quality control (QC) consisted of 1,125 individuals. The mean age was 14.28 years (s.d. = 1.71), and 508 participants (45.15%) were males.

### Depression

Lifetime and past year MDD and MDE measures were attained via participant reports on the World Health Organization Composite International Diagnostic Interview (CIDI) and based on DSM-IV criteria (American Psychiatric Association, 1994). The clinical validity of CIDI in relation to standardized clinical assessments has been supported (Kessler et al., 2009). Notably, the clinical algorithm included in WMH-CIDI-A can differentiate between those cases whose depression is best explained by an MDD diagnosis and those whose depression is related to other comorbid disorders, such as bipolar disorder. In the present study, we applied a hierarchical diagnostic algorithm that classified participants as MDE cases if they met the criteria for a depressive episode, regardless of comorbid disorders, but classified participants as MDD cases only if their depressive symptoms were better explained by MDD than by a different diagnosis, such as bipolar disorder.

### Adversity

Information about 12 psychosocial adversities experienced during childhood was collected. Example adversities assessed include child neglect, abuse, parental psychopathology, death of a parent, or family economic challenges. These adversities were evaluated from the childhood and posttraumatic stress disorder sections of the WMH-CIDI-A as described elsewhere (Benjet et al., 2011). The selection and scoring of these measures are the same as those developed for the World Mental Health Survey (Green et al., 2010). An adversity sum score (ADV) reflects the number of adverse situations a given participant endorsed.

### Genotyping and genotype data processing

Participants were asked to provide a mouthwash sample following clinical interviews. Two thousand four hundred fifty-eight participants (81.79% of the sample) agreed to provide a biospecimen. DNA was extracted using the Puregene® protocol. Genotyping was performed at the Broad Institute of Harvard and MIT for a subset (∼1,300) of samples with high DNA-yield, using the Illumina Global Screening Array (GSA). Pre-imputation quality control consisted in: i) removal of variants with high missingness (geno 0.05), low minor allele frequency (MAF < 0.01) and with evidence of deviance from Hardy-Weinberg equilibrium (p_HWE_< 1e-6) ii) removal of individuals with genotype missingness > 5% (n = 1) and individuals with extreme heterozygosity (±3 standard deviations from the mean; n = 16). The final pre-imputation QCed dataset consisted of 1,152 participants and 406,862 variants.

### Genotype imputation

Before imputation, genotype data strand, allele assignment (reference and alternate), and positions were compared and adjusted to the 1000Genomes reference panel using the McCarthy Group software tools (https://www.well.ox.ac.uk/~wrayner/tools/). Imputation was carried out using the Michigan Imputation Server (https://imputationserver.sph.umich.edu/) using Minimac4 and the 1000g-phase-3-v5 (hg19) full reference panel (admixed population). Imputed genotype data were transformed to plink dosage format using DosageConvertor. A principal component plot illustrating the genetic makeup of the sample can be found in the supplementary materials (**Supplementary Figure 1**). As shown in Figure 1, the ancestral contributions to our sample from Mexico City (CDMX) were predominantly Native American (0.71, s.d. = 0.14; AMR) and European (0.26, s.d. = 0.12; EUR), and to a lesser extent, African (0.03, s.d. = 0.02; AFR).

**Fig. 1.**
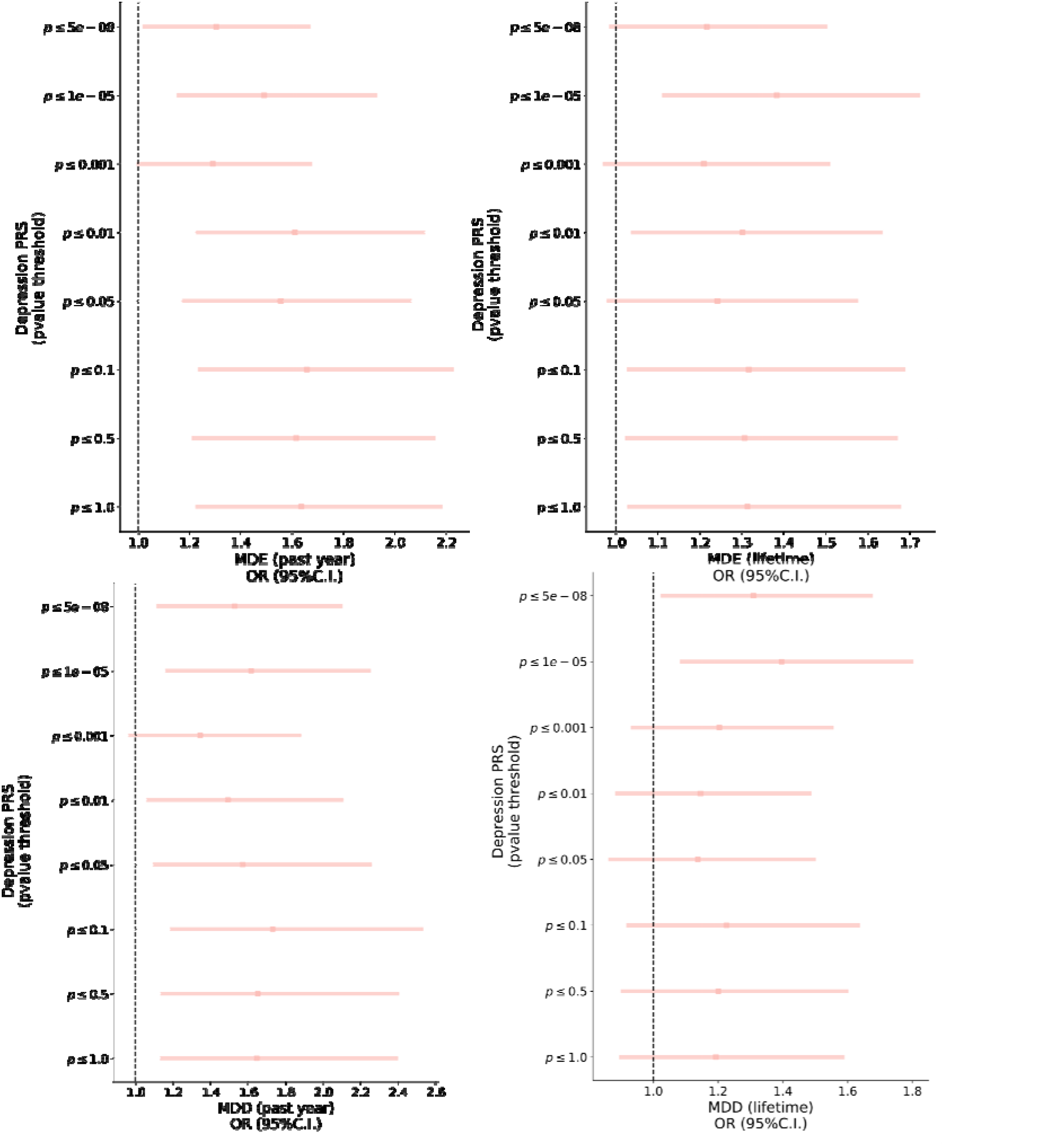
Depression PRS and phenotypic depression associations. Forest plot depicting the results of a logistic regression testing for the association between depression PRS and depression phenotypes. X- axis depicts the phenotype odds ratio (OR) per PRS standard deviation increase. Y-axis depicts the SNP inclusion criteria (p value threshold)

### Depression GWAS Discovery Sample

PRS were created based on the largest GWAS meta-analysis of depression to date, which included 807,553 individuals (246,363 cases and 561,190 controls) of European ancestry (Howard et al., 2019). The cohorts included in the meta-analysis were the UK Biobank, 23andMe, Inc., and the Psychiatric Genomics Consortium.^11^ Summary statistics for the UK Biobank and PGC cohorts were obtained from the online repository)https://datashare.is.ed.ac.uk/handle/10283/3203), whereas summary statistics for the 23andMe cohort were obtained under an agreement with 23andMe that protects the privacy of the 23andMe participants (http://research.23andme.com/collaborate/#publication for more information).

### Polygenic risk scoring

PRS were calculated for the target sample using the depression summary statistics referenced above. Variants for PRS estimation were further QCed by excluding indel, strand ambiguous- and low (R2< 0.6) imputation quality variants. The most significant independent SNPs were selected using a conservative clumping procedure in PLINK 1.98 (Chang et al., 2015). (p1 = 1, p2 = 1, r2 = 0.1, kb = 10000) to correct for inflation arising from linkage disequilibrium (LD). Eight PRS were calculated using different p-value thresholds (p< 5×10–8, p< 1×10–5, p< 0.001, p< 0.01, p< 0.05, p< 0.1, p< 0.5, p< 1) as criteria for SNP inclusion on the PRS. PRS were calculated by multiplying the effect size of a given SNP by the imputed allelic dosage of the effect allele present in an individual. Finally, the SNP dosage effects were summed across all loci per individual. Associations between depression PRS and phenotypes of interest (e.g., MDD in the past 12 months) were assessed using a logistic regression analysis implemented by the *statsmodels* module in *python*. All of the associations were covaried for the effects of age, sex, and the first 20 genetic principal components to account for population stratification; the standardized (z-transformed) PRS were included in the analyses as a predictor of interest.

### Multiple testing correction

In this study, we tested the association between four depression categories (i.e., MDD diagnosis and MDE for both lifetime and past year) and eight PRS for depression. Since a high correlation is expected between diagnoses, but also between PRS as they are calculated using overlapping sets of SNPs, we employed the matrix spectral decomposition approach to control for multiple testing (Nyholt, 2004). Briefly, the Eigen decomposition of the correlation matrix of the eight PRS and the four phenotypes was used to estimate the number of effective independent variables (V_eff_ = 6). We, therefore, set the experiment-wide significance threshold at p< 0.0085 to keep a type I error rate <= 5%.

### G x E interaction

We first assessed the association between lifetime adversity and the different depression diagnoses using logistic regression models. We then tested whether the interaction between the depression PRS and environmental effects (i.e., an adversity score [ADV]) were associated with depression diagnoses. Interaction analyses were only conducted if the outcome of interest was significantly associated with adversity. Due to the skewness of the adversity score, we defined high adversity as reporting more than three types of adversity. This approach has already been successfully used in this (Cruz-Fuentes et al., 2014) and other samples (Kessler and Bromet, 2013). Finally, the sample was split into low, medium, and high genetic risk for depression based on PRS tertile. Two models, one including the effect of G x E (e.g., depression PRS * ADV) and another one restricting this effect to be zero, were fitted using maximum likelihood estimation. A likelihood ratio test (LRT) statistic between the full and restricted models was calculated. The significance of the LRT was estimated using a chi-square test with one degree of freedom. These analyses were performed in *python* using the *statsmodels* and *scipy* modules.

### Ethics approval and informed consent

This study was conducted in accordance with the ethical principles of the Declaration of Helsinki as revised 1989 and was approved by the Ethics and Scientific Committees of the National Institute of Psychiatry “Ramón de la Fuente Muñiz” (INP-RFM) in Mexico City. Interviewers gave a verbal and written explanation of the study and obtained informed consent from the parent or legal guardian and the assent of the adolescent.

## Results

The prevalence of both MDD and MDE was higher for the female subgroup (6–13%) compared to the male subgroup (2–6%). As expected, the diagnosis with the highest prevalence was lifetime history of any major depressive episode (**Table 1**). Results from the primary analyses are presented below.

**Table 1.** Effect sizes of Depression PRS on past year and lifetime MDD diagnosis and MDE.

| Diagnosis | % cases | OR (95% CI) | P-value | Partial R<sup>2</sup> |
| --- | --- | --- | --- | --- |
| **MDD past year** | **3.97%** | **1.73 (1.18-2.53)** | **4.7e-3** | **0.028** |
| **MDE past year** | **6.75%** | **1.65 (1.23-2.23)** | **9.0e-4** | **0.028** |
| MDD lifetime | 6.31% | 1.40 (1.08-1.80) | 0.0104 | 0.016 |
| **MDE lifetime** | **9.33%** | **1.38 (1.11-1.72)** | **3.9e-3** | **0.017** |

Note. Bolded cells reflect significant findings after correcting for multiple testing.

### Association between depression PRS and MDD/MDE

All the effect sizes of depression PRS on depression diagnoses were positive (**Table 1 and Figure 1**). Depression PRS showed statistically significant associations with MDE (both lifetime and in the past 12 months) and with MDD in the past 12 months. Nonetheless, the associations between depression PRS and lifetime MDD did not show a consistent statistically significant association across the different p-value thresholds (**Figure 1**) and failed to reach statistical significance after correcting for multiple testing (**Table 1**).

### Interactions between adversity and depression PRS

Past year MDD and lifetime MDE showed statistically significant (p< 0.0123 Bonferroni adjusted threshold) associations with adversity (**Table 2**); thus, interaction analyses were conducted considering these outcomes (past year MDD and lifetime MDE). Despite a visual trend suggesting interactions between some of the depression PRS and adversity predicting MDD in the past year (**Supplementary Figure 2)** and lifetime MDE (**Supplementary Figure 3**), the associations between the depression PRS and adversity interactions did not reach statistical significance for any of the PRS p-value SNP inclusion thresholds (**Supplementary Table 1**).

**Table 2.** GxE interaction results in predicting past year and lifetime MDD diagnosis and MDE.

| **Phenotype** | **PRS**<br>**P-value**<br>**cutoff** | **OR (95% CI)** | **LRT<sup>a</sup>** | **P-value** |
| --- | --- | --- | --- | --- |
| MDD past year | 5.00E-08 | 0.53 (0.24-1.18) | 17.42 | 0.6852 |
|  | 1.00E-05 | 0.59 (0.25-1.43) | 16.63 | 0.7331 |
|  | 0.001 | 0.70 (0.31-1.59) | 15.74 | 0.7843 |
|  | 0.01 | 0.49 (0.22-1.10) | 18.48 | 0.6183 |
|  | 0.05 | 0.66 (0.28-1.54) | 15.89 | 0.7758 |
|  | 0.1 | 0.89 (0.42-1.92) | 15.70 | 0.7861 |
|  | 0.5 | 1.29 (0.57-2.93) | 15.58 | 0.7929 |
|  | 1 | 1.25 (0.55-2.83) | 15.63 | 0.7902 |
| MDE lifetime | 5.00E-08 | 0.53 (0.24-1.18) | 17.42 | 0.6852 |
|  | 1.00E-05 | 0.59 (0.25-1.43) | 16.63 | 0.7331 |
|  | 0.001 | 0.70 (0.31-1.59) | 15.74 | 0.7843 |
|  | 0.01 | 0.49 (0.22-1.10) | 18.48 | 0.6183 |
|  | 0.05 | 0.66 (0.28-1.54) | 15.89 | 0.7758 |
|  | 0.1 | 0.89 (0.42-1.92) | 15.70 | 0.7861 |
|  | 0.5 | 1.29 (0.57-2.93) | 15.58 | 0.7929 |
|  | 1 | 1.25 (0.55-2.83) | 15.63 | 0.7902 |

Note. GxE interaction analyses were only conducted for the significant associations between depression outcomes and adversity.
<sup>a</sup>LRT = Likelihood ratio test.

## Discussion

Depression is a debilitating illness that is associated with significant psychosocial, physical, and occupational impairments (Chiu et al., 2018; Costello et al., 2003), with recent work indicating significant incidence and recurrence among Mexican youth (Benjet et al., 2019). While advances in GWAS have facilitated the study of novel genetic loci associated with this disorder (Howard et al., 2019), most of this research has only considered European ancestry populations. Determining whether findings from these studies apply to samples of Mexican youth is critical, given differences in population history, allele frequencies, and linkage disequilibrium that may influence the strength of the genetic associations observed. Building on the limited work that has examined the relationship between depression polygenic risk, adversity, and phenotypic depression in non-European samples (Peterson et al., 2017), we examined whether depression polygenic risk was associated with phenotypic depression, and whether adversity exposure influenced the association between depression polygenic risk and depression diagnoses in a diverse, representative sample of adolescents from Mexico City.

We observed significant positive associations between depression PRS and phenotypic depression. This finding is consistent with previous work linking higher polygenic loading for MDD with elevated depressive symptoms during adolescence in European-ancestry populations (Kwong et al., 2019, 2020; Rice et al., 2019). Notably, the depression PRS accounted for between 1.5%-2.5% of the variance in past year MDD and MDE, paralleling the amount of variance explained in European samples (1.5%-3.2%) (Howard et al., 2019). The similar amount of variance accounted for in phenotypic depression is surprising as there is often a net attenuation in the variance accounted for when there is a mismatch between the ancestry of the GWAS compared to the target sample (Martin et al., 2018). There are, however, several explanations to explain the comparable amount of variance accounted for in our study compared to the discovery sample. First, as noted previously, the discovery GWAS sample size was very large and well powered, which may have contributed to our ability to detect significant associations in our sample. Second, relative to early-onset childhood depression, whereby environmental factors often have a greater influence on symptoms, adolescent-onset depression seems to be more heavily influenced by genetic predispositions (Rice et al., 2002). For example, the heritability of depression has been shown to vary by age such that in childhood and young adulthood, heritability estimates peak in adolescence (*h*^2^ = .17) (Sallis et al., 2017). Greater genetic influences on depressive symptoms may also be due to gene-environment correlations such that during adolescence, youth have greater independence from caregivers and may seek out environments (e.g., peer selection) that reflect their negative world view (Jaffee and Price, 2012).

Consistent with previous studies that have indicated a positive association between adversity exposure and depressive symptoms among Mexican and Latino adolescents (Benjet et al., 2019; Jaschek et al., 2016), we found that higher levels of adversity were linked to a greater likelihood of experiencing MDD and MDE in one’s lifetime and the past year. However, we did not observe significant interactions between depression PRS and adversity with phenotypic MDD or MDE. These findings are consistent with work that suggests that stressful life events did not influence the association between depression polygenic risk and depression among European ancestry adults (Mullins et al., 2016). One explanation for the lack of significant interaction effects may be that most people in the sample reported relatively low levels of adversity (only 79 participants out of ∼1200 reported facing more than three types of adversity). Thus, we may have been unable to detect G x E effects given the low frequency of adversity at the higher end of the adversity continuum.

There are some limitations to this study that must be acknowledged. The data are cross-sectional, which prevented us from establishing causality among the constructs. For example, while it is possible that higher levels of adversity may contribute to the development of negative cognitive schemas that play a role in depression onset (Wickrama et al., 2016), it is also possible that experiencing a depressive episode might result in more negative perceptions of past events. Another question that we were not able to address here is the possibility that adverse life events may result in epigenetic modifications, which may exacerbate depression susceptibility. Future work should consider collecting biospecimens across several time points, in addition to multiple reports of adversity and depressive symptoms, to understand the biological routes through which experiences become embedded.

Despite these limitations, our work represents an essential first step in attempting to delineate (a) whether genetic loci identified in the largest GWAS to date on depressive symptoms are related to phenotypic depressive symptoms, and (b) whether adverse life events influenced these relationships in an underrepresented population in genomics research. The PRS approach holds immense promise for informing precision science and personalized medicine initiatives. Already, there has been an increased use of PRS in clinical settings, with PRS being used to inform risk of coronary artery disease, type 2 diabetes, and breast cancer, with prediction results that are comparable to health conditions that are a result of monogenic mutations (Khera et al., 2018). When it comes to complex psychiatric phenotypes, such as depression, the clinical usefulness of PRS is still in its infancy. However, one can conceive of a future where the consideration of PRS, in combination with other clinical risk factors, are used to anticipate and possibly mitigate risk for phenotypic depression onset. Before such a future can be realized, PRS must be accurate and generalizable to all populations to not further contribute to health inequities (Martin et al., 2018). Efforts aimed at attenuating heath disparities, such as the recruitment of diverse ancestry populations in gene discovery efforts, as well as research that validates the use of PRS in multiancestry samples, such as that presented here, is imperative.

## Data Availability

Summary statistics for the Depression PGC cohort are available in the online repository (https://datashare.is.ed.ac.uk/handle/10283/3203). Access to summary statistics for the 23andMe cohort require scientist collaborators to enter an agreement with 23andMe that protects the privacy of the 23andMe participants (http://research.23andme.com/collaborate/#publication for more information).
Access to individual data of adolescent participants is restricted by applicable policies and regulations that protect their privacy. Potential academic collaborators are encouraged to contact the authors to discuss potential collaboration. Code used as part of this study is available from the authors upon reasonable request.

## Declarations of interest

No conflicts of interest to declare.

## Author contributions

J.A.R. and M.E.R. conceived the present PRS study. C.B. and C.S.C-F designed and directed the epidemiological and genetic components of the adolescent mental health population survey, respectively, including all data and biospecimen collection. Genetic data QC and imputation were conducted by A.I.C. with support from L.M-K. Phenotypic data was cleaned and handled by G.A.M-L and E.M. The first draft was led by J.A.R. (introduction and discussion), G.A.M-L (methods) and A.I.C. (results). J.S. contributed to the conceptualization of the manuscript and assisted in editing. M.E.R. coordinated the project, and all authors provided intellectual input and feedback on the manuscript.

## Acknowledgements

The authors would like to thank all staff involved in the epidemiological project for their support and participation, and to the youth that agreed to participate in the study and donate biological samples. The original survey was carried out in conjunction with the World Health Organization World Mental Health (WMH) Survey Initiative. We thank the WMH staff for assistance with instrumentation and fieldwork. We also thank the research participants and employees of 23andMe for their contribution to the original depression discovery GWAS. This study was supported by grant SEP 2004 COI 46594 (PI C.S. Cruz-Fuentes), grant CB 60678 and support from fund 0196 of Mexico’s National Institute of Psychiatry (PI C. Benjet). Funds were employed to pay salaries of interviewers, reagents and laboratory material. We thank Prof. Nick Martin from QIMR Berghofer for useful comments and feedback on preliminary versions of this work. A.I.C. is supported by a UQ Research Training Scholarship from The University of Queensland (UQ). M.E.R. thanks the support of a fellowship from Australia’s National Health and Medical Research Council (APP1102821).

## Supplementary material

**Supplementary Table 1.** Association between adversity score and lifetime and past year MDD diagnosis and MDE.

| Diagnosis | OR (95% CI) | P-value |
| --- | --- | --- |
| **MDD past year** | 1.20 (0.96-1.50) | 0.1080 |
| **MDE past year** | **1.38 (1.17-1.63)** | **0.0001** |
| MDD lifetime | 1.13 (0.95-1.35) | 0.1639 |
| **MDE lifetime** | **1.29 (1.12-1.49)** | **0.0005** |

Note. Bolded cells reflect significant findings after correcting for multiple testing

**Supplementary Fig. 1.**
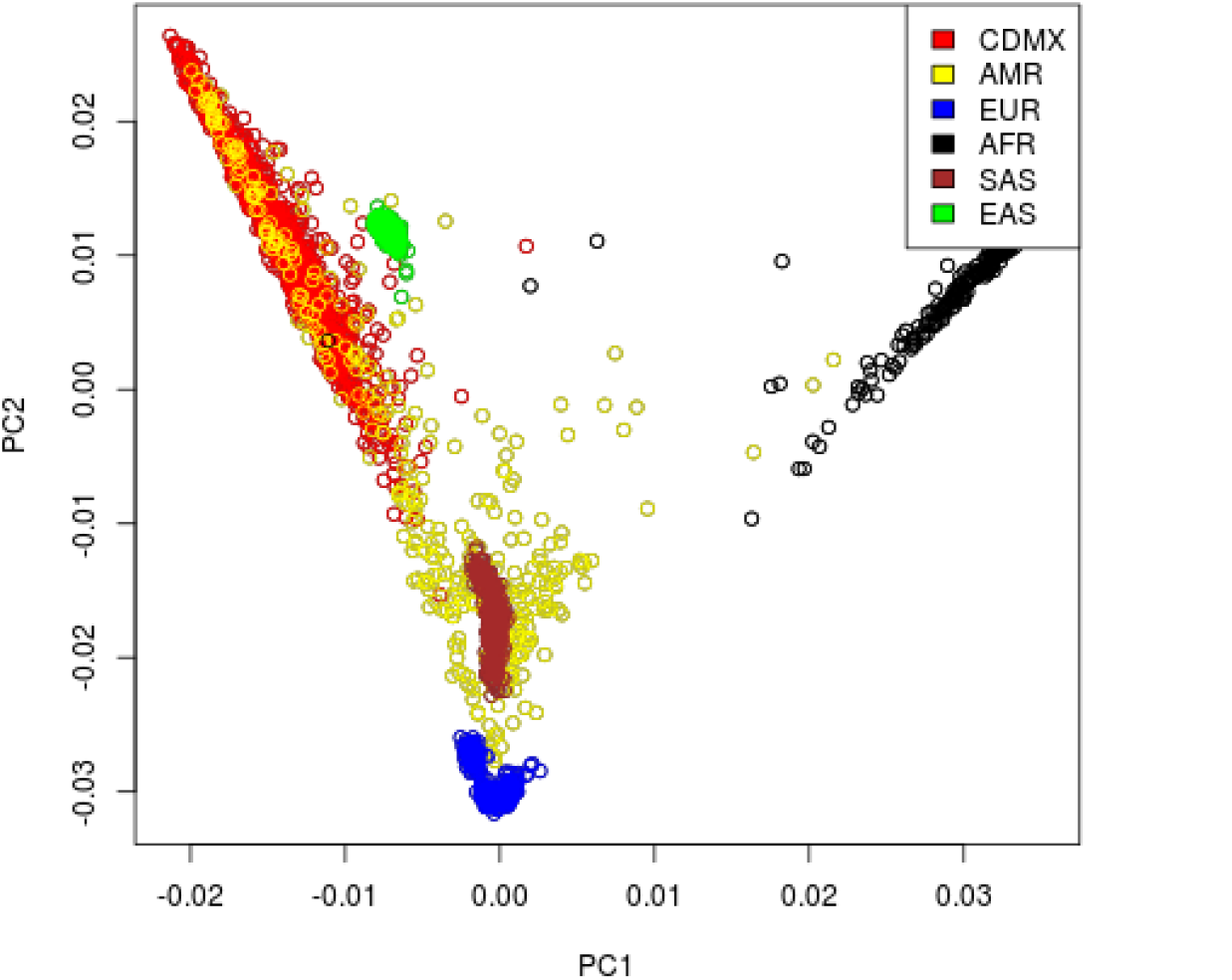
Ancestry Principal Component Plot. Scatterplot of the first two ancestry principal components reflects the admixed composition of the Mexico City (CDMX) sample. 1000 Genomes Project Individuals from a ncestral continental of tal n; populations are provided for reference (AMR = Native American; EUR = European; AFR = African; SAS = South Asian; EAS = East Asian).

**Supplementary Fig. 2.**
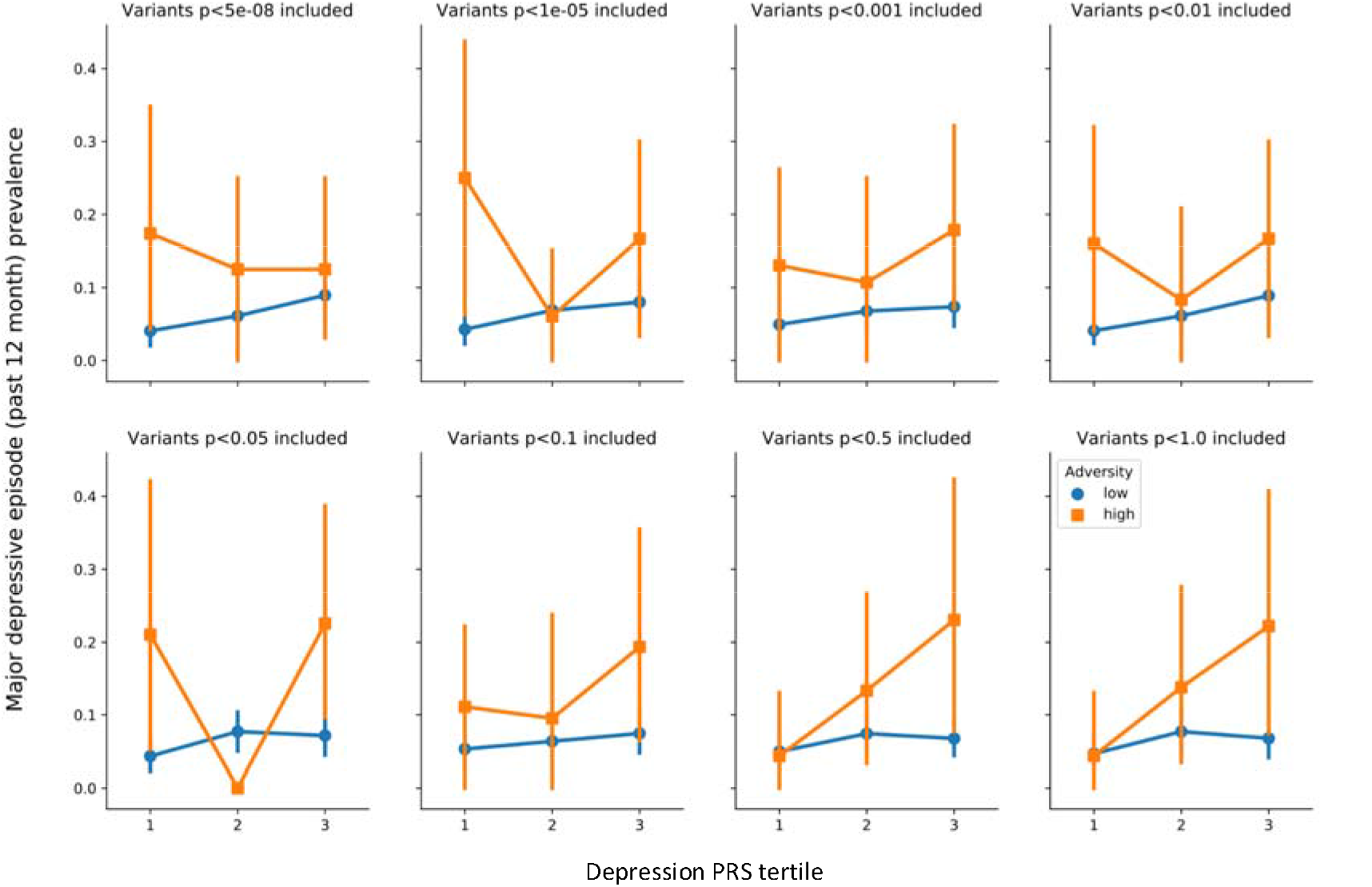
GxE interaction for past year MDD. Plots showing the depression PRS * adversity score interaction. The x-axis shows the PRS divided in tertiles as a proxy for low, medium and high genetic risk for MDD. The y-axis shows the prevalence of major depressive episode in the past year. The sample is split into low risk (blue circles) and high risk (orange squares). An interaction would imply a statistically significant change in the slope between both lines.

**Supplementary Fig. 3.**
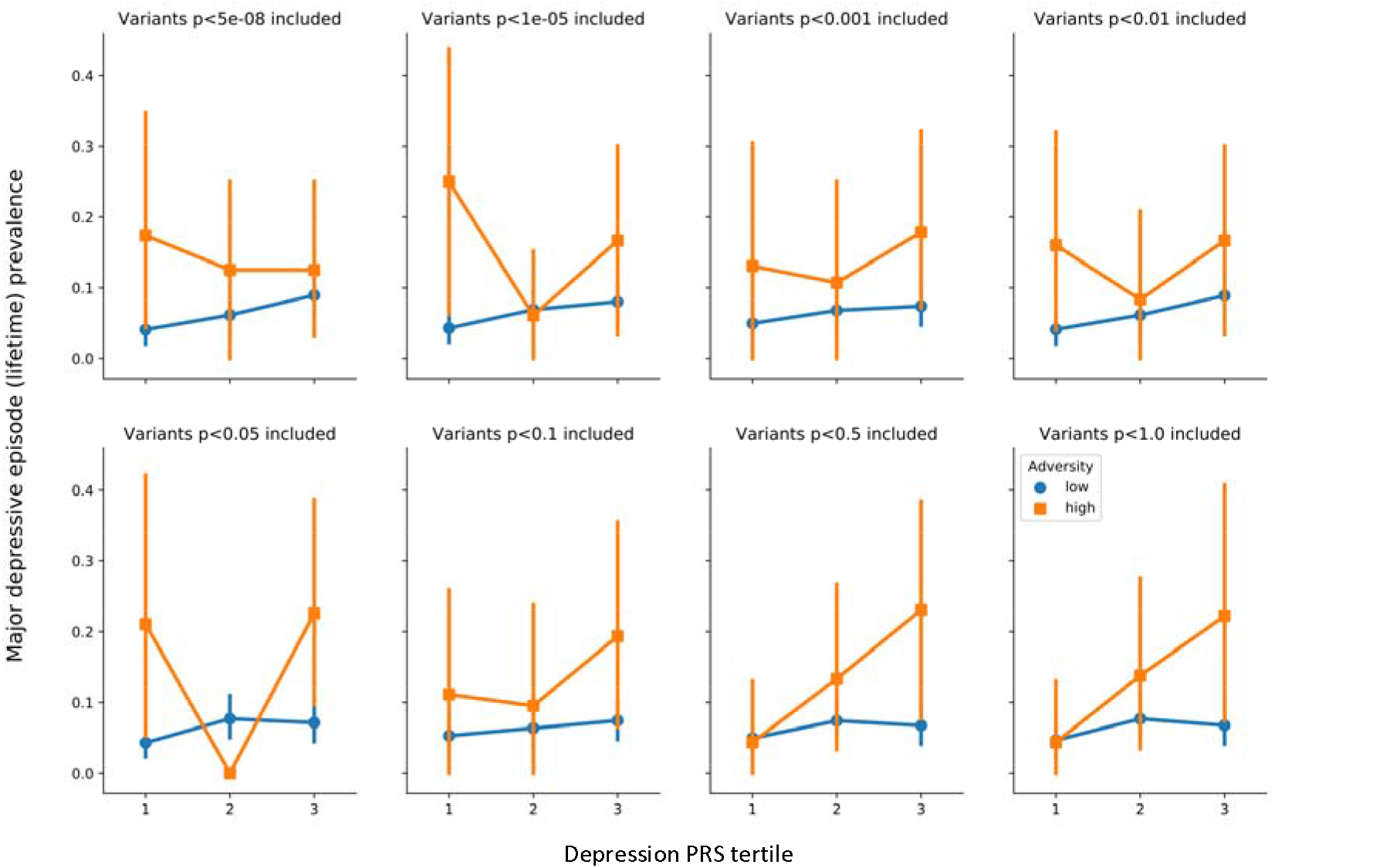
GxE interaction for lifetime MDE. Plots showing the interaction between depression PRS * adversity score. The x-axis shows the PRS divided in tertiles as a proxy for low, medium and high genetic risk for MDD. The y-axis shows the prevalence of major depressive episode in the past year. The sample is split into low risk (blue circles) and high risk (orange squares). An interaction would imply a statistically significant change in the slope between both lines.

## Notes

### Competing Interest Statement

The authors have declared no competing interest.

### Funding Statement

This study was supported by grant SEP-2004-COI-46594 (PI C.S. Cruz-Fuentes), grant CB-2006-01-60678 and support from fund 0196 of Mexico's National Institute of Psychiatry (PI C. Benjet). Funds were employed to pay salaries of interviewers, reagents and laboratory material.

### Author Declarations

Ethics approval and informed consent. This study was conducted in accordance with the ethical principles of the Declaration of Helsinki as revised 1989 and was approved by the Ethics and Scientific Committees of the National Institute of Psychiatry "Ramon de la Fuente Muniz" (INP-RFM) in Mexico City. Interviewers gave a verbal and written explanation of the study and obtained informed consent from the parent or legal guardian and the assent of the adolescent.

